# Interim BNT162b2 LP.8.1-adapted COVID-19 Vaccine Effectiveness among Non-Immunocompromised Adults ≥65 years of age in California and Louisiana, August–December 2025

**DOI:** 10.64898/2026.07.27.26359050

**Authors:** Laura D. Zambrano, Tiange Yu, Jazmine Mateus, Xin Zhao, Kathleen M. Andersen, Srinivas R. Valluri, Burcu Karakuzu Ikizler, Rajeev M. Nepal, Adam MacNeil, Hannah R. Volkman

## Abstract

Evidence on vaccine effectiveness (VE) of 2025-26 COVID-19 vaccines is limited. We estimated VE of BNT162b2 LP.8.1-adapted vaccine among non-immunocompromised adults ≥65 years through December 2025 using linked claims and immunization registry data from two U.S. states. VE against COVID-19-related ED/UC encounters was 48% (95% CI:19, 66).

## Background

COVID-19 remains a major cause of morbidity in older U.S. adults. In October 2024, the LP.8.1 variant (JN.1 lineage) emerged and became predominant by April 2025 [1]. Given distinct antigenic characteristics that diverged from the previously circulating KP.2 variant, the U.S. Food and Drug Administration authorized updated LP.8.1-adapted monovalent messenger RNA (mRNA) COVID-19 vaccines on August 27, 2025 [2].

Estimates of vaccine effectiveness (VE) of LP.8.1-adapted COVID-19 vaccines during the 2025-26 viral respiratory season are limited. In this analysis, we estimate interim VE of the BNT162b2 LP.8.1-adapted COVID-19 vaccine through December 2025 among non-immunocompromised adults ≥65 years against COVID-19-related emergency department or urgent care (ED/UC) encounters, using linked claims and immunization registry data from two U.S. states.

## Methods

We conducted a retrospective cohort study among non-immunocompromised adults ≥65 years of age in California and Louisiana who were enrollees of health insurance and pharmacy plans captured in HealthVerity using previously established methods [3]. California and Louisiana are states with complete reporting of COVID-19 immunizations to registries. This data source includes state registry immunization data, which is linked via tokenization to data from medical (diagnostic and procedural) and pharmacy claims among approximately 30% of the population in each state; claims represent those submitted by commercial insurance, Medicare, and Medicaid Managed Care plans [4]. Enrolled adults were followed from August 27, 2025 (day of FDA authorization for the updated LP.8.1-adapted formulation) to December 31, 2025.

Eligible individuals entered the cohort on August 27, 2025 (index date) if they had 1) ≥1 year of continuous California or Louisiana residency and 2) ≥1 year of continuous medical and pharmacy enrollment (allowing ≤30-day gaps to account for renewal gaps) to enable ascertainment of comorbidities and previous healthcare utilization. Individuals were excluded if they had an immunocompromising condition or were receiving immunosuppressive therapy within one year prior to the index date, had discrepancies or missingness in sex or year of birth between the claims database and the state immunization registries, had a diagnosis of COVID-19 (*International Classification of Diseases, Tenth Revision, Clinical Modification* [ICD-10-CM] code U07.1) within 90 days before index, or had a COVID-19 vaccine administered within 90 days before index (**Supp Table 1**).

### Patient Consent Statement

Given the use of deidentified data, this study was deemed exempt from institutional review board review by the Sterling Institutional Review Board (ID #13433) and did not include factors necessitating patient consent.

### Exposure

Vaccination was considered a time-varying exposure, whereby adults were considered exposed 7 days after receiving a BNT162b2 LP-8.1-adapted vaccine dose, as recorded through either administrative claims or the state immunization registry. Adults were considered unexposed if they did not receive a 2025-26 COVID-19 vaccine of any type, indicated by the absence of any 2025-26 COVID-19 vaccine in both administrative claims and state immunization registry. Unexposed persons contributed unexposed person-time throughout follow-up, whereas vaccinated persons contributed unexposed person-time until the date of vaccination, after which they contributed exposed person-time from 7 days post-vaccination through the end of follow-up.

To augment vaccine verification, administrative claims data were linked to state immunization registry data in California and Louisiana via tokenization [7] while maintaining de-identification to ensure Health Insurance Portability and Accountability Act (HIPAA) compliance [8]. California and Louisiana mandate reporting to their state immunization registries within 24 hours and one week of COVID-19 vaccine administration, respectively.

### Outcomes

The primary outcome was COVID-19-related ED/UC encounters defined as those with an accompanying U07.1 ICD-10 diagnosis code. The outcome was evaluated as events occurring per 100,000 person-months at risk among exposed and unexposed cohorts.

### Statistical analysis

Standardized mean differences (SMD) were used to evaluate potential differences between groups, with SMD >0.1 indicating imbalance. Cox proportional hazards regression was used to model adjusted hazard ratios (aHR) with 95% confidence intervals (CI) for the occurrence of each outcome. Vaccine effectiveness was modeled as [(1 – aHR) X 100], with models adjusted for sex (male or female), state of residence (California or Louisiana), insurance payor (Medicaid, Medicare, or commercial), presence of any condition(s) indicating CDC-defined high risk for severe COVID-19 in the year prior to index (any or none) [9], wellness visit in the one year before index (yes or no), influenza vaccination in the year before index (yes/no), outpatient visits within 180 days before index (yes or no), ED visits within 180 days before index (yes or no), and medically attended COVID-19 in the 91 to 365 days before index (yes or no). Models used the Efron method for ties in time-to-event between individuals. Individuals were followed from index to event or were censored on December 31, 2025, disenrollment from a medical or pharmacy plan, date of receipt of a non-BNT162b2 2025-26 vaccine, or receipt of a second 2025-26 COVID-19 vaccine dose of any kind (**Supp Table 2**). Additional stratified analyses examined VE among adults ≥65 years of age with ≥1 high-risk underlying medical condition and those with no high-risk underlying medical conditions as defined by CDC [9]. All analyses were performed using SAS V9.4 (SAS Institute) and R V4.5.0 (R Foundation for Statistical Computing). Reporting from this study followed the Enhancing the QUAlity and Transparency Of health Research (EQUATOR) guidelines for the REporting of studies Conducted using Observational Routinely collected health Data statement for PharmacoEpidemiology (RECORD-PE) [5]. The study protocol was posted on clinicaltrials.gov (NCT06923137) prior to initiation of all analyses [6].

## Results

After applying exclusion criteria, 1,369,461 adults were included, of whom 1,245,970 (91%) resided in California and 123,491 (9%) resided in Louisiana. The median age was 72 years (interquartile range [IQR]: 68, 78), 55% of all participants were female, and 14%, 30%, and 55% were enrolled in commercial/private insurance, Medicaid, or Medicare plans, respectively. During the study period, 104,476 (7.6%) received the BNT162b2 LP.8.1 vaccine, including 97,318 (7.8%) in California and 7,158 (5.8%) in Louisiana (**Table**). There were 21 COVID-19-related ED/UC encounters among those who received the BNT162b2 LP.8.1 vaccine and 1,630 COVID-19-related ED/UC encounters among those who did not receive a 2025-26 COVID-19 vaccine of any kind (**Figure**).

**Table.** Demographic characteristics of adults ≥65 years who received the BNT162b2 LP.8.1-adapted formulation during the 2025-26 season and those who did not receive any 2025-26 COVID-19 vaccine, California and Louisiana, August 27 to December 31, 2025

|  |  | Exposure Status |  |  |
| --- | --- | --- | --- | --- |
|  | *Overall, n (%)*<br>*(N=1,369,461)* | *Did not receive any*<br>*2025-26 COVID-19*<br>*vaccine, n (%)*<br>*(N= 1,264,985)* | *Received BNT162b2*<br>*LP.8.1 COVID-19*<br>*vaccine, n (%)*<br>*(N=104,476)* |  |
| Characteristic |  |  |  | *SMD* |
| *State of residence* |  |  |  |  |
| California | 1,245,970 (91.0) | 1,148,652 (90.8) | 97,318 (93.1) | 0.09 |
| Louisiana | 123,491 (9.0) | 116,333 (9.2) | 7,158 (6.9) |  |
| *Age (years)* |  |  |  |  |
| Median (IQR) | 72 (68, 78) | 72 (68, 78) | 73 (69, 78) |  |
| *Sex* |  |  |  |  |
| Female | 746,688 (54.5) | 690,739 (54.6) | 55,949 (53.6) | 0.02 |
| Male | 622,773 (45.5) | 574,246 (45.4) | 48,527 (46.4) |  |
| *Insurance type* |  |  |  |  |
| Commercial/private | 197,389 (14.4) | 176,198 (13.9) | 21,191 (20.3) | 0.37 |
| Medicaid | 415,401 (30.3) | 398,083 (31.5) | 17,318 (16.6) |  |
| Medicare | 747,856 (54.6) | 683,501 (54.0) | 64,355 (61.6) |  |
| Unknown | 8,815 (0.6) | 7,203 (0.6) | 1,612 (1.5) |  |
| *Influenza vaccination in prior year* |  |  |  |  |
| Documented receipt | 974,625 (71.2) | 931,087 (73.6) | 43,538 (41.7) | 0.68 |
| No documented receipt | 394,836 (28.8) | 333,898 (26.4) | 60,938 (58.3) |  |
| *Wellness visit in prior year* |  |  |  |  |
| At least one | 522,523 (38.2) | 475,306 (37.6) | 47,217 (45.2) | 0.16 |
| None | 846,938 (61.8) | 789,679 (62.4) | 57,259 (54.8) |  |
| *Outpatient visit in prior 180 days* |  |  |  |  |
| At least one | 810,219 (59.2) | 740,187 (58.5) | 70,032 (67.0) | 0.18 |
| None | 559,242 (40.8) | 524,798 (41.5) | 34,444 (33.0) |  |
| *ED visit in prior 180 days* |  |  |  |  |
| At least one | 131,017 (9.6) | 122,057 (9.6) | 8,960 (8.6) | 0.04 |
| None | 1,238,444 (90.4) | 1,142,928 (90.4) | 95,516 (91.4) |  |
| *COVID-19 diagnosis in prior 91 - 365 days* |  |  |  |  |
| At least one | 17,448 (1.3) | 15,710 (1.2) | 1,738 (1.7) | 0.04 |
| None | 1,352,013 (98.7) | 1,249,275 (98.8) | 102,738 (98.3) |  |
| *Any history of a high-risk underlying medical condition associated with severe COVID-19<sup>a</sup>* |  |  |  |  |
| No high-risk condition | 354,943 (25.9) | 328,460 (26.0) | 26,483 (25.3) | 0.01 |
| ≥1 high-risk condition | 1,014,518 (74.1) | 936,525 (74.0) | 77,993 (74.7) |  |
| Asthma | 135,163 (9.9) | 123,318 (9.7) | 11,845 (11.3) | 0.05 |
| Cerebrovascular disease | 243,439 (17.8) | 226,135 (17.9) | 17,304 (16.6) | 0.03 |
| Chronic heart disease<sup>b</sup> | 374,421 (27.3) | 346,583 (27.4) | 27,838 (26.6) | 0.02 |
| Chronic kidney disease<sup>c</sup> | 26,688 (1.9) | 25,245 (2.0) | 1,443 (1.4) | 0.05 |
| Chronic liver disease<sup>d</sup> | 145,112 (10.6) | 134,511 (10.6) | 10,601 (10.1) | 0.02 |
| Chronic lung disease<sup>e</sup> | 240,104 (17.5) | 222,843 (17.6) | 17,261 (16.5) | 0.03 |
| Cystic fibrosis | 310 (0.0) | 291 (0.0) | 19 (0.0) | <0.01 |
| Diabetes mellitus | 506,310 (37.0) | 470,006 (37.2) | 36,304 (34.7) | 0.05 |
| Disabilities | 539,277 (39.4) | 496,044 (39.2) | 43,233 (41.4) | 0.04 |
| Hematologic malignancies | 19,982 (1.5) | 18,342 (1.5) | 1,640 (1.6) | <0.01 |
| Immunocompromising conditions<sup>f</sup> | 84,401 (6.2) | 77,854 (6.2) | 6,547 (6.3) | <0.01 |
| Mental health disorders | 314,457 (23.0) | 290,420 (23.0) | 24,037 (23.0) | <0.01 |
| Neurologic disorders | 110,226 (8.0) | 103,325 (8.2) | 6,901 (6.6) | 0.06 |
| Obesity (BMI ≥30 kg/m<sup>2</sup>) | 398,935 (29.1) | 369,765 (29.2) | 29,170 (27.9) | 0.03 |
| Current or former smoker | 315,959 (23.1) | 292,335 (23.1) | 23,624 (22.6) | 0.01 |
| Tuberculosis | 3,676 (0.3) | 3,447 (0.3) | 229 (0.2) | 0.01 |

Abbreviations: ED = emergency department; BMI = body mass index; IQR = interquartile range; kg/m<sup>2</sup> = kilograms per meters squared; N/n = number; SMD = standardized mean difference
<sup>a</sup> High-risk conditions were present if they aligned with CDC guidance (https://www.cdc.gov/covid/hcp/clinical-care/underlying-conditions.html)
<sup>b</sup> Including heart failure, coronary artery disease, or cardiomyopathy.
<sup>c</sup> Requiring dialysis
<sup>d</sup> Limited to cirrhosis, non-alcoholic fatty liver disease, alcoholic liver disease, and autoimmune hepatitis
<sup>e</sup> Limited to bronchiectasis, chronic obstructive pulmonary disease, interstitial lung disease, pulmonary embolism, and pulmonary hypertension
<sup>f</sup> Adults were excluded from the analysis if any immunocompromising condition was reported within the period of the one-year period before index. The immunocompromising conditions reported in this table reflect any documented in individuals' medical history.

**Figure.**
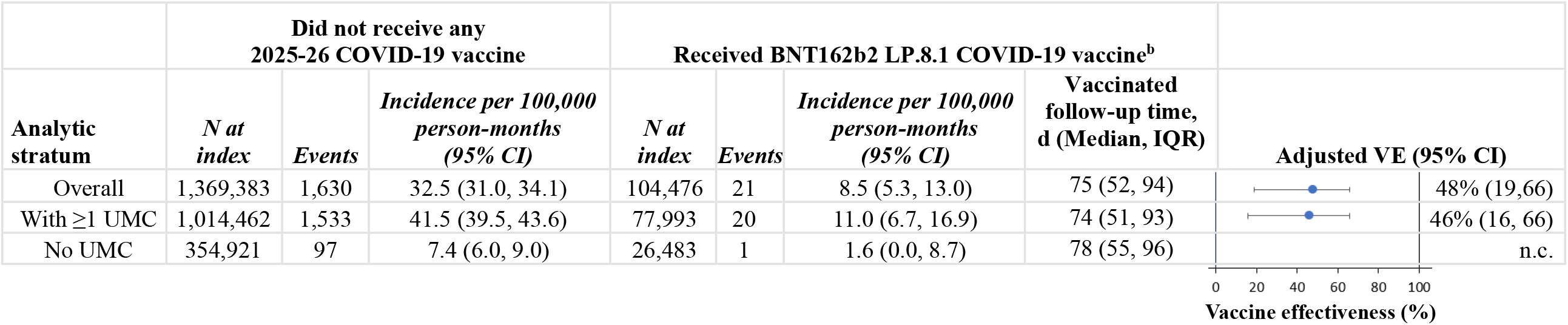
Estimates of BNT162b2 LP.8.1-adapted COVID-19 vaccine effectiveness^a^ against laboratory-confirmed COVID-19-associated healthcare encounters among adults ≥65 years, California and Louisiana, August 27, 2025 to December 31, 2025 Abbreviations: CI =confidence interval; d = days; ED = emergency department; HR = hazard ratio; IQR = interquartile range; N = number; n.c. = not calculable due to insufficient power and event count in the vaccinated group; UC = urgent care; UMC = high-risk underlying medical condition; VE =vaccine effectiveness; ^a^ VE was calculated through Cox proportional hazards regression with vaccination as a time-varying exposure. Adjusted models included the following time-fixed covariates: Sex (male or female), state of residence (California or Louisiana), insurance payor (Medicaid, Medicare, or commercial), presence of any condition(s) indicating CDC-defined high risk for severe COVID-19 in the year prior to index (any or none), wellness visit in the year prior to index (yes/no), influenza vaccination in the year before index (yes/no), any outpatient visit 180 days before index (yes/no), emergency department visits 180 days before index (yes/no), and medically attended COVID-19 in the 91 to 365 days before index (yes/no) ^b^ Seven days were required from date of vaccine dose to be considered exposed/vaccinated. Adults with an event from days from 0 to 6 days post-vaccination were not counted in either the exposed or unexposed group. **Alt text:** Forest plot depicting point estimate for vaccine effectiveness (blue dot) and 95% confidence intervals (error bars) for adults aged ≥65 years, stratified by the presence of at least one high-risk underlying medical condition.

CDC-defined high-risk underlying medical conditions were enumerated in 1,014,518 (74.1%) adults, with the most common being qualifying disabilities (39.4%), diabetes mellitus (type 1 or 2) (37.0%), and obesity (29.1%). The prevalence of ≥1 high-risk underlying medical condition was similar for exposed and unexposed persons (74.7% among those who received the BNT162b2 LP.8.1 vaccine vs 74.0% among those who did not receive a 2025-26 COVID-19 vaccine of any kind, SMD=0.01). The frequency of each underlying medical condition was similar (SMD<0.06 for all conditions) by vaccination status (**Table**).

Overall VE of the BNT162b2 LP.8.1-adapted vaccine against COVID-19-related ED/UC encounters among adults ≥65 years of age was 48% (95% CI: 19, 66) at a median of 75 (IQR 52, 94) days since dose, and 46% (95% CI: 16, 66) at a median of 74 (IQR 51, 93) days since dose among adults ≥65 years of age with ≥1 high-risk underlying medical condition (**Figure**). VE could not be calculated among adults ≥65 years without high-risk underlying medical conditions due to insufficient power. Within this group, one person of 26,483 who received a BNT162b2 LP.8.1 vaccine had a COVID-19-related ED/UC encounter (incidence rate [IR]: 1.6, 95% CI: 0.04, 8.7 encounters per 100,000 person-months), compared with 97 of 354,921 adults who did not receive any 2025-26 COVID-19 vaccine (IR: 7.4, 95% CI: 6.0, 9.0 encounters per 100,000 person-months). The cumulative incidence of COVID-19-related ED/UC encounters according to time-varying vaccination status reflects significantly lower incidence of COVID-19-related ED/UC encounters over the follow-up period among adults who received the BNT162b2 LP.8.1-adapted COVID-19 vaccine compared with those who received no 2025-26 COVID-19 vaccine (**Supp Figure**).

## Discussion

In this retrospective cohort study conducted between August 27 and December 31, 2025 among nearly 1.4 million non-immunocompromised adults ≥65 years of age in California and Louisiana, receipt of the BNT162b2 LP.8.1-adapted vaccine provided significant protection against ED/UC encounters associated with COVID-19 compared with those who did not receive any 2025-26 COVID-19 vaccine during this period. These results demonstrate the added benefit of the BNT162b2 LP.8.1-adapted COVID-19 vaccine in the setting of high population immunity and support routine administration of this vaccine in older adults.

These findings are consistent with emerging evidence supporting the effectiveness of 2025-26 COVID-19 vaccines. Three test-negative case-control studies from the United States and Canada reported VE estimates of between 48% and 57% against ED/UC encounters or medically attended COVID-19, including COVID-19-related ED/UC encounters [10–12]. The biological plausibility of these findings is supported by pseudoneutralization studies demonstrating preserved activity against both XFG and LP.8.1 variants which circulated in the U.S. during the study [13]. Notably, the consistency of VE estimates across seasons[14] and convergence of VE estimates across North American studies with varying healthcare systems, analytic platforms, and using both test-negative and retrospective cohort designs underscores the robustness, reproducibility, and validity of observed LP.8.1 vaccine effectiveness across all studies.

This analysis has several limitations. First, it was restricted to California and Louisiana, the only states with linked state immunization registries within the HealthVerity environment, which may limit generalizability to the broader U.S. population. Second, COVID-19 cases were identified using the ICD-10-CM diagnosis code U07.1, and incidental COVID-19 is challenging to rule out for non-hospitalized adults using claims data alone. Third, although adults with medically attended COVID-19 within 90 days of the index date were excluded, we could not rule out all recent infections, such as those that were subclinical in nature. Fourth, although vaccination status was modeled as a time-varying exposure to reduce immortal time bias, survivor bias and residual confounding may persist in retrospective observational vaccine effectiveness studies [15]. Finally, these findings reflect only the first four months of the 2025-26 respiratory virus season, precluding assessment of the effects of waning protection or viral evolution over time.

The BNT162b2 LP.8.1-adapted COVID-19 vaccine conferred significant added protection against COVID-19-related ED/UC encounters among non-immunocompromised adults ≥65 years of age. These findings support routine administration of COVID-19 vaccine formulations targeting currently circulating variants in older adults.

## Data Availability

Aggregate data produced in the present study are available upon reasonable request to the authors

## Disclosures

Competing interests: Pfizer Inc. funded this study. LDZ, KMA, SRV, BKI, RMN, AM, and HRV are employees of Pfizer Inc. LDZ, KMA, SRV, BKI, RMN, AM, and HRV hold stocks or stock options in Pfizer Inc. TY, JM, and XZ are employees of Genesis Research Services, which received funding from Pfizer in connection with the development of this manuscript.

## Funding support

This study was sponsored by Pfizer Inc.

## Role of the funder/sponsor

All authors participated, as employees of or contractors to Pfizer Inc., in the design and conduct of the study; collection, management, analysis, and interpretation of the data; preparation, review and approval of the manuscript; and decision to submit the manuscript for publication.

## Notes

### Clinical Protocols

https://clinicaltrials.gov/study/NCT06923137

### Author Declarations

Given the use of deidentified data, this study was deemed exempt from institutional review board review by the Sterling Institutional Review Board (ID #13433).

