## Supplemental Material for "Interim BNT162b2 LP.8.1-adapted COVID-19 Vaccine Effectiveness among Non-Immunocompromised Adults ≥65 years of age in California and Louisiana, August–December 2025"

**Supplemental Table 1.** Attrition steps yielding the final analytic dataset for vaccine effectiveness analyses of BNT162b2 LP.8.1-adapted vaccine in non-immunocompromised adults ≥65 years, California and Louisiana, August 27, 2025 to December 31, 2025

| | Overall | | State of Residence | | | |
|---|---|---|---|---|---|---|
| | | | California | | Louisiana | |
| **Cohort Selection Steps** [a] | ***N*** | ***%*** | ***N*** | ***%*** | ***N*** | ***%*** |
| 1. All unique patients who were California or Louisiana residents in the database as of **August 27, 2025** (index date) and born in **1960** or earlier | 4,372,889 | | 3,718,601 | | 655,278 | |
| 2. and at least one year of pharmacy and medical enrollment prior to index date. A gap of up to 30 days is allowed. | 2,157,582 | 49.34% | 1,930,841 | 51.92% | 227,349 | 34.70% |
| 3. and at least one year of continuous single state residency in California or Louisiana prior to index | 2,074,779 | 47.45% | 1,858,691 | 49.98% | 216,088 | 32.98% |
| 4. and is not currently immunocompromised (using up to 1 year lookback) at the time of index | 1,479,345 | 33.83% | 1,345,298 | 36.18% | 134,047 | 20.46% |
| 5. and without null values or discrepancies in sex and/or year of birth between claims and California/Louisiana immunization registry datasets | 1,403,174 | 32.09% | 1,278,059 | 34.37% | 125,115 | 19.09% |
| 6. and without a diagnosis of COVID-19 in any setting ≤ 90 days prior to index | 1,395,191 | 31.91% | 1,270,784 | 34.17% | 124,407 | 18.99% |
| 7. and without receipt of COVID-19 vaccine < 90 days prior to index | 1,369,461 | 31.32% | 1,245,970 | 33.51% | 123,491 | 18.85% |

[a] Initial sum of population does not equal total, as initial enrollee identification included persons dually enrolled in both California and Louisiana. Attrition does not account for single-state enrollment until Step 3.

**Supplemental Table 2.** Distribution of censoring events by vaccination status among non-immunocompromised adults ≥65 years of age

| Censoring event | Did not receive any 2025-26 COVID-19 vaccine of before end of follow-up<br>n (%) | Received BNT162b2 LP.8.1 COVID-19 vaccine before end of follow-up<br>n (%) |
|---|---|---|
| **Total patients by vaccination status** | 1,264,985[a] | 104,476 |
| **Outcome (COVID-19 diagnosis in ED/UC)** | 1,630 | 21 |
| **Events** | | |
| Medical/pharmacy disenrollment | 432,462 (34.2) | 31,471 (30.1) |
| Receipt of second dose | - | 0 |
| Receipt of non-BNT162b2 vaccine | 80,453 (6.4) | 0 |
| End of follow-up (12/31/2025) | 973,047 (76.9) | 89,797 (85.9) |

Abbreviations: ED/UC = emergency department/urgent care; n = number

[a] Individuals who remained unexposed through December 31, 2025, after removing those who contributed unexposed person-time up the point of achieving fully vaccinated status 7 days after receipt of their BNT162b2 LP.8.1-adapted COVID-19 vaccine.

**Supplemental Table 3.** Distribution of medically attended COVID-19 in the ED/UC setting over time, by presence of high-risk underlying medical conditions, among adults ≥65 years of age.

| Month of first COVID-19 diagnosis | With high-risk underlying medical condition (N=1,553) | Without high-risk underlying medical condition (N=98) |
|---|---|---|
| | **n (%)** | **n (%)** |
| August 2025 | 173 (11.1) | 8 (8.2) |
| September 2025 | 914 (58.9) | 57 (58.2) |
| October 2025 | 262 (16.9) | 21 (21.4) |
| November 2025 | 109 (7.0) | 8 (8.2) |
| December 2025 | 95 (6.1) | 4 (4.1) |

Abbreviations: ED/UC = emergency department / urgent care; N/n = number

**Supplemental Figure.** Cumulative incidence with 95% confidence limits of COVID-19-associated emergency department and urgent care encounters, by vaccination status among non-immunocompromised adults ≥65 years, California and Louisiana, August 27, 2025 to December 31, 2025

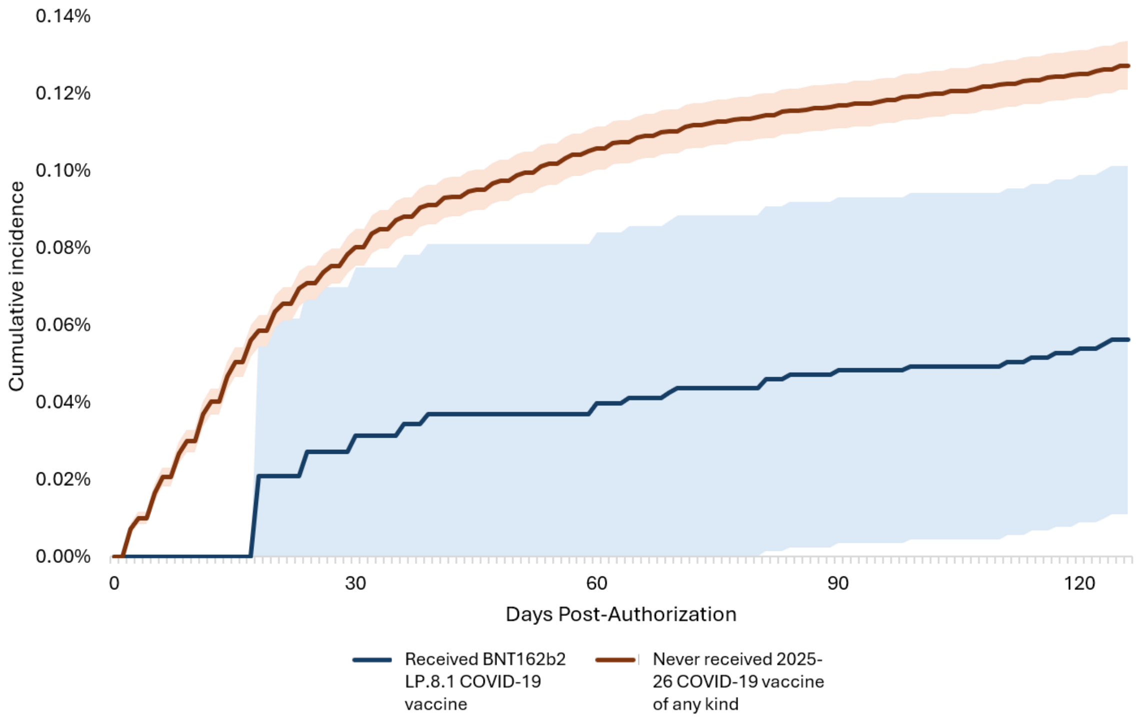

**Alt text:** Plot of cumulative incidence (%) of COVID-19-related ED/UC encounters (y-axis) on each day after August 27, 2025, the date of FDA authorization for LP.8.1-adapted mRNA vaccines (x-axis). Those who received the LP.8.1-adapted BNT162b2 vaccine are shown in blue, while those who never received a 2025-26 COVID-19 vaccine of any kind are shown in orange. All estimates are accompanied by shading representing upper and lower 95% confidence limits.
